# Caregivers’ burden of care during emergency department care transitions among older adults: a mixed methods cohort study

**DOI:** 10.1101/2024.07.16.24309597

**Authors:** Nathalie Germain, Estephanie Jémus-Gonzalez, Vanessa Couture, Émilie Côté, Michèle Morin, Annie Toulouse-Fournier, Laetitia Bert, Raphaëlle Giguère, Samir Sinha, Nadia Sourial, Lucas B. Chartier, Holly O. Witteman, France Légaré, Rawane Samb, Stéphane Turcotte, Sam Chandavong, Lyna Abrougui, Joanie Robitaille, Patrick M. Archambault, the Network of Canadian Emergency Researchers

## Abstract

**Objective:** Improving care transitions for older adults can reduce emergency department (ED) revisits, and the strain placed upon caregivers. We analyzed whether caregivers felt a change in burden following a care transition, and what may be improved to reduce it.

**Methods:** This mixed-methods observational study nested within LEARNING WISDOM included caregivers of older patients who experienced an ED care transition. Burden was collected with the brief Zarit Burden Interview (ZBI-12), and caregivers commented on the care transition. A qualitative coding scheme of patient care transitions was created to reflect themes important to caregivers. Comments were randomly analyzed until saturation and themes were extracted from the data. We followed both the SRQR and STROBE checklists.

**Results:** Comments from 581 caregivers (mean age (SD) 64.5 (12.3), 68% women) caring for patients (mean age (SD) 77.2 (7.54), 48% women) were analyzed. Caregivers overwhelmingly reported dissatisfaction and unmet service expectations, particularly with home care and domestic help. Communication and follow-up from the ED emerged as an area for improvement. Caregivers who reported an increased level of burden following a care recipient’s care transition had significantly higher ZBI scores than caregivers who self-reported stable burden levels, but not improved burden levels.

**Conclusion:** Caregivers with increasing, stable, and improved levels of subjective burden all reported areas for improvement in the care transition process. Themes centering on the capacity to live at home most frequently and may represent serious challenges to caregivers. Addressing these challenges could improve both caregiver burden and care transitions.

**Key points:**

1. We analyzed caregivers’ thoughts about emergency department care transitions using both qualitative and quantitative tools.
2. Caregivers reported dissatisfaction and unmet service expectations with home care, domestic help, and coordinating follow-ups.
3. Variance in self-reported subjective caregiver burden corresponds to Zarit Burden Interview (ZBI) scores.

## Background

Populations around the world are undergoing a significant demographic shift, marked by the steady growth of older adults as a segment of society, and the shrinking of the available workforce of those who care for older people [1]. Within this context, unpaid caregivers, primarily family members, spouses, and adult children, are being increasingly tasked to fill in the gap [2]. Most caregiving tasks from activities of daily living to medical procedures are now the responsibility of family caregivers, with healthcare system services complementing the care significantly often being provided by caregivers [3]. Caregivers help older adults adhere to care and medication schedules, remain in their homes, and avoid premature institutionalization, while also playing a critical role in the passage of patients between levels of health care and across care settings (e.g., from an emergency department (ED) to the community).

While caregiving can be rewarding, it entails substantial commitment and responsibilities, which can exact a toll on caregivers both physically and emotionally. This confluence of challenges is commonly referred to as caregiver burden. Burden can arise from the intensity and duration of caregiving responsibilities, a lack of support or coping mechanisms, and the condition and behavior of the care recipient (CR) [4]. High levels of caregiver burden may predispose caregivers to burnout and health issues, thereby impairing their capacity to provide effective care [5]. The health of patients may deteriorate if their caregiver is overwhelmed or unable to deliver adequate care, resulting in poor outpatient clinical outcomes and an increase in avoidable ED revisits [6,7].

Cooperation between caregivers, healthcare providers, and patients is crucial to the success of care transitions, but challenges still abound [8,9]. Caregivers have previously reported that discharge plans are often drawn by healthcare providers that require the caregiver—without consulting the caregiver as to the feasibility of the plan [8,9]. Patients may also decline crucial professional services like bathing or administering medications, preferring having their caregiver perform these tasks. This may be to the detriment of the caregiver, who may not be comfortable taking on that role [8,9]. Clinicians often face ambiguity as to when is the best time to involve a family caregiver in discussing levels of care, or how to adjudicate when there is a disagreement [10–12]. Consulting with and discussing care plans with caregivers has the potential to enhance patient and caregiver satisfaction with communication, better navigate available resources, and to alleviate the discomfort clinicians feel in discussions of transitioning or changing levels of care [13].

The purpose of this mixed-methods descriptive study is to analyze in tandem ZBI scores, and the contents of comments collected from the caregivers of older patients having experienced a care transition.

## Method

### Study design and context

This study was nested within the longitudinal cohort study of an integrated health research project of **[ANON]** (CISSS-**[ANON]**): LEARNING WISDOM (*Supporting the Creation of a LEARNing INteGrated Health System to Mobilize Context-adapted Knowledge with a Wiki Platform to Improve the Transitions of Frail Seniors From Hospitals and Emergency Departments to the cOMmunity*) [16]. The LEARNING WISDOM cohort included older adults and their caregivers who underwent a transition of care following a visit to one of four EDs in the CISSS-**[ANON]** between January 2019 and December 2021. The CISSS-**[ANON]** is an integrated health organization consisting of four acute care hospitals: A, B, C and D. A is a university teaching hospital receiving more than 70,000 annual ED visits while the other three rural sites each receive around 30,000 visits.

The protocol for this study was approved by the CISSS-**[ANON]** Ethics Review Committee (project #2018-462, 2018-007). We adhered to the Standards for Reporting Qualitative Research (SRQR) [17] guidelines for the assessment of qualitative outcomes and employed The Strengthening the Reporting of Observational Studies in Epidemiology (STROBE) Statement [18] to report the quantitative outcomes.

### Participants

LEARNING WISDOM included consenting patients aged 65 years or older, who had been discharged back to the community from the ED observation unit. Patients only seen in the ambulatory care section of the ED, admitted to hospital, transferred to another hospital, or transferred to a long-term care center were excluded. Patients and their caregivers had to understand and speak French. For the full duration of the study recruitment period, at each participating hospital, a list of eligible discharged patients was generated each day. Patient phone numbers were selected using a computer-generated daily randomized list and patients were contacted to participate. No additional eligibility criteria were added for this specific study.

### Data collection

Using a deductive approach with an a-priori coding scheme developed in a previous study of patient comments [15], we designed this mixed-methods descriptive study to analyze data collected directly from the caregivers of older patients having experienced a care transition. As part of a continuous quality improvement project led by the CISSS-**[ANON]**, patients were contacted by telephone between 24h to 7 days after ED discharge, to administer the three-item Care Transitions Measure (CTM-3) [19,20]. Patients were then invited to participate in a more in-depth research interview in the following days.

During this second call, patients consented based on the Nova Scotia Criteria [21] and as such were required to summarize—in their own words—their understanding of the study to participate. Participating patients were then asked if they consented to have their caregivers contacted by the research team. Demographic characteristics for patients and caregivers were collected using a structured interview, while some patient characteristics (e.g., comorbidities), were collected with chart review.

Informed consent was then also obtained for all contacted caregivers, who were then administered the Zarit Burden Interview (ZBI). The ZBI is the most widely used instrument measuring caregiver burden [22]. The reliability of scores on the ZBI measured by internal consistency (Cronbach’s alpha) is high, between 0.70–0.93 [23–25]. This short version of the ZBI consists of 12 items in two domains, and consists of two constructs, including role strain (items 1–9) and personal strain (10–12). Each question is scored by frequency in a five-point Likert scale (0 to 4): 0 for never, 1 for rarely, 2 for sometimes, 3 for quite often, and 4 for all the time. The scores are then summed into an overall indication of burden (range 0–48). For caregivers of patients with dementia, Hébert provided cut-offs < 3 as low burden, 3–8 as moderate, 9–18 as high, and > 18 as severe [14].

Caregivers also answered two open-ended questions in as much or as little detail as they wished. Translated from the original French, the first one (Question A) was: “*In your opinion, has there been a change in the burden of care following your [CR]’s departure from the emergency department*?”. The second (Question B) was: “*In your opinion, what could be improved to reduce the burden of care for your [CR]?*”. Research professionals recorded critical elements of each patient’s response with important verbatim excerpts in REDCap (Research Electronic Data Capture) [26,27]. These professionals (research nurses, and two PhD psychology students) were trained by the research team and authorized by the Director of Nursing and the Professional Services Director to perform data collection.

### Developing the framework for content analysis

We used a mixed inductive and deductive approach for content analysis. We first used a hypothetical-deductive framework, seeking to amend an existing model of patient experiences of care transitions [15] to capture and systematically analyze the perspectives of caregivers as told through open-ended response data [28,29]. The original coding framework included 4 main themes and 19 sub-themes (See Appendix D) and involved noting when a sub-theme appeared in a comment (1 for affirmative, 0 for no mention) in addition to its emotional valence. Emotional valence reflects the extent to which a comment reads as positive or negative in its statement. We used a quantifiable metric scaling system in which we rate the emotional valence of the comment: 0 negative, 1 positive, and 2 neutral. Importantly, we coded the absence of a requested service or item as negative because we argue that unmet needs are negative in emotional valence.

Inter-rater reliability was established by coding 40 randomly selected comments in parallel. The resulting reliability coefficient was high (Krippendorff’s Alpha: 0.90) [30,31]. Disagreements were resolved by discussion between the two coders (NG and EJG) and the principal investigator (PA). The analysis was performed by two female evaluators from different scientific backgrounds (NG, an MSc student in epidemiology with training in mixed methods, and EJG, an MD student with research and clinical experience) and supervised by an experienced clinician researcher with expertise in qualitative analyses (PA) [32,33].

The original four main themes that emerged when analyzing patient comments regarding a recent transition of care were *Care in the emergency department*, *Conditions of stay*, *Independent living at home*, and *Discharge* [15]. It was planned that after coding comments using this patient-centered coding scheme, we would turn to an inductive-deductive approach by shifting the model from patient-centered to caregiver-centered. We then returned to a hypothetical-deductive approach and systematically applied an amended model of caregiver experiences to the open-ended response data.

Two coders (NG and EJG) performed content analysis until saturation, stopping when additional comments did not reveal new themes [34]. Each individually coded 30 randomly selected comments per hospital (selection without replacement) [34], then additional randomly selected comments in rounds of 10. Saturation was achieved when coding 2 consecutive rounds of 10 without the emergence of a new theme per hospital. Inter-rater reliability of binary coded data was calculated with Cohen’s Kappa [35].

### Statistical and visual analyses

An a-priori power analysis was conducted for LEARNING WISDOM and is described elsewhere [16]. No a-priori power analysis was conducted for the analyses in this article. For caregivers included in content analysis, we conducted a binomial test of the proportion of each self-reported burden change category versus chance. If the groups were distributed randomly, and there was no pattern of changes in subjective burden following a care transition, we would expect each category contain 25% of participants.

To corroborate scores on the ZBI with caregiver reports as to how their level of burden may have increased. We conducted a one-way analysis of variance (ANOVA) of subjective change in burden on ZBI score. The assumption of homogeneity of variance was violated (Levene’s test *F*(3, 577) = 3.18, *p* = .023), so we conducted the ANOVA with a Brown-Forsythe correction.

We also conducted a one-way analysis of covariance (ANCOVA) to determine if a difference on ZBI scores exists as a function of their self-reported change in subjective burden while controlling for age and comorbidities.

For comments containing two or more themes, we used the *quanteda* package in R (Quantitative Analysis of Textual Data) [36] to visualize and organize concurrent themes within caregiver comments using a series of co-occurrence network plots.

## Results

The total LEARNING WISDOM cohort included 5,016 participating patients (Figure 1). Patients (*n* = 1819) allowed the research team to contact their caregiver, and 410 caregivers were excluded or declined to participate, leaving 1,409 patient-caregiver dyads. Of these, 778 caregivers provided open-response comments to Question A (Appendix A, French version). Of these caregivers, 752 responded to Question B.

**Figure 1.**
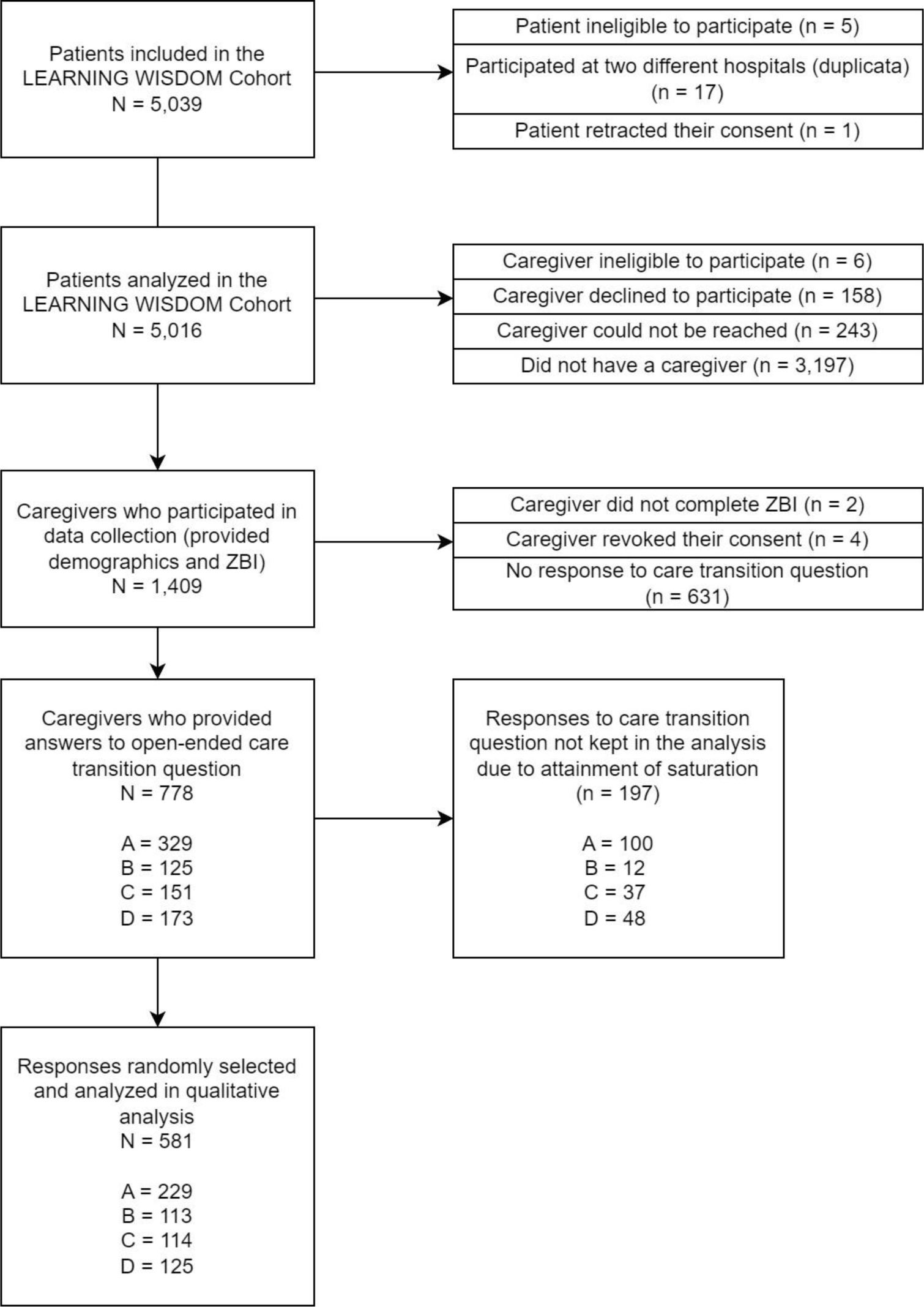
Flowchart describing the recruitment of CR patients and their caregivers.

### Qualitative Results

#### Analysis of themes within the content analysis

Of 778 caregiver responses to what could be improved to reduce the burden of care for a CR, 581 were analyzed: 229 from A, 125 from B, 114 from C and 113 from D. Demographic characteristics of 581 caregivers and their CR are found in Table 1, stratified by self-reported change in burden. Of these, 235 of these comments mentioned at least one main theme and 328 caregiver comments did not contain any themes. Concerning the overall valence of these 253 comments, 60 mentions were positive (23.7%), 33 neutral (13%), and 160 negative (63.2%).

**Table 1.** Demographic characteristics of care recipient (CR) patients and caregivers stratified by the caregiver’s self-reported change in burden.

|  | **No**<br>**comment**<br>**(N=26)**<br>**4.4%** | **Burden**<br>**increased**<br>**(N=138)**<br>**23.7%** | **Burden**<br>**unchanged**<br>**(N=374)**<br>**64.3%** | **Burden**<br>**improved**<br>**(N=43)**<br>**7.4%** | **Overall**<br>**(N=581)** |
| --- | --- | --- | --- | --- | --- |
| **Hospital** |  |  |  |  |  |
| A | 10 (38.5%) | 66 (47.8%) | 137 (36.6%) | 16 (37.2%) | 229 (39.4%) |
| B | 3 (11.5%) | 21 (15.2%) | 79 (21.1%) | 10 (23.3%) | 113 (19.4%) |
| C | 10 (38.5%) | 25 (18.1%) | 72 (19.3%) | 7 (16.3%) | 114 (19.6%) |
| D | 3 (11.5%) | 26 (18.8%) | 86 (23.0%) | 10 (23.3%) | 125 (21.5%) |
| **Patient age** |  |  |  |  |  |
| Mean (SD) | 80.1 (8.34) | 78.4 (7.33) | 76.5 (7.34) | 77.7 (8.70) | 77.2 (7.54) |
| **Patient gender** |  |  |  |  |  |
| Man | 12 (46.2%) | 65 (47.1%) | 198 (52.9%) | 27 (62.8%) | 302 (52.0%) |
| Woman | 14 (53.8%) | 73 (52.9%) | 176 (47.1%) | 16 (37.2%) | 279 (48.0%) |
| **Arrival at the ED** |  |  |  |  |  |
| Ambulance | 12 (46.2%) | 83 (60.1%) | 184 (49.2%) | 20 (46.5%) | 299 (51.5%) |
| Walk-in | 14 (53.8%) | 55 (39.9%) | 190 (50.8%) | 23 (53.5%) | 282 (48.5%) |
| **Canadian Triage**<br>**Acuity Scale (CTAS)** |  |  |  |  |  |
| 1 | 0 (0%) | 0 (0%) | 1 (0.3%) | 0 (0%) | 1 (0.2%) |
| 2 | 2 (7.7%) | 16 (11.6%) | 44 (11.8%) | 9 (20.9%) | 71 (12.2%) |
| 3 | 17 (65.4%) | 65 (47.1%) | 198 (52.9%) | 17 (39.5%) | 297 (51.1%) |
| 4 | 7 (26.9%) | 51 (37.0%) | 123 (32.9%) | 13 (30.2%) | 194 (33.4%) |
| 5 | 0 (0%) | 6 (4.3%) | 8 (2.1%) | 4 (9.3%) | 18 (3.1%) |
| **Time on stretcher (Hours)** |  |  |  |  |  |
| Mean (SD) | 10.3 (7.36) | 12.1 (8.57) | 10.5 (8.31) | 14.3 (7.85) | 11.2 (8.36) |
| **Charlson Comorbidity Index Score** |  |  |  |  |  |
| Mean (SD) | 6.12 (2.58) | 5.04 (1.73) | 5.06 (2.09) | 5.42 (2.30) | 5.13 (2.06) |
| **Covid-19 Wave at time of ED visit (Québec)\*** |  |  |  |  |  |
| Pre-pandemic | 18 (69.2%) | 107 (77.5%) | 259 (69.3%) | 26 (60.5%) | 410 (70.6%) |
| Wave 1 | 7 (26.9%) | 29 (21.0%) | 111 (29.7%) | 16 (37.2%) | 163 (28.1%) |
| Between the end of wave 1 and wave 2 | 1 (3.8%) | 2 (1.4%) | 4 (1.1%) | 1 (2.3%) | 8 (1.4%) |
| **Have a family physician** |  |  |  |  |  |
| Yes | 24 (92.3%) | 131 (94.9%) | 352 (94.1%) | 39 (90.7%) | 546 (94.0%) |
| No | 2 (7.7%) | 7 (5.1%) | 22 (5.9%) | 4 (9.3%) | 35 (6.0%) |
| **Can quickly get an appointment with family physician if needed** |  |  |  |  |  |
| Yes | 18 (69.2%) | 77 (55.8%) | 239 (63.9%) | 28 (65.1%) | 362 (62.3%) |
| No | 8 (30.8%) | 61 (44.2%) | 135 (36.1%) | 15 (34.9%) | 219 (37.7%) |
| **Have access to transport** |  |  |  |  |  |
| Yes | 23 (88.5%) | 127 (92.0%) | 347 (92.8%) | 41 (95.3%) | 538 (92.6%) |
| No | 3 (11.5%) | 11 (8.0%) | 27 (7.2%) | 2 (4.7%) | 43 (7.4%) |
| **People in social circle** |  |  |  |  |  |
| Mean (SD) | 4.31 (5.14) | 3.30 (2.53) | 4.07 (3.57) | 3.72 (2.33) | 3.87 (3.37) |
| **First language** |  |  |  |  |  |
| French | 26 (100%) | 138 (100%) | 373 (99.7%) | 43 (100%) | 580 (99.8%) |
| Missing | 0 (0%) | 0 (0%) | 1 (0.3%) | 0 (0%) | 1 (0.2%) |
| **Patient ethnicity** |  |  |  |  |  |
| Caucasian | 26 (100%) | 138 (100%) | 374 (100%) | 41 (95.3%) | 579 (99.7%) |
| Missing | 0 (0%) | 0 (0%) | 0 (0%) | 2 (4.7%) | 2 (0.3%) |
| **Patient highest level of education** |  |  |  |  |  |
| Primary school | 11 (42.3%) | 60 (43.5%) | 155 (41.4%) | 20 (46.5%) | 246 (42.3%) |
| High school (DES) | 6 (23.1%) | 45 (32.6%) | 96 (25.7%) | 11 (25.6%) | 158 (27.2%) |
| College (DEC) | 4 (15.4%) | 12 (8.7%) | 40 (10.7%) | 3 (7.0%) | 59 (10.2%) |
| Vocational studies (DEP) | 2 (7.7%) | 11 (8.0%) | 34 (9.1%) | 3 (7.0%) | 50 (8.6%) |
| University studies | 3 (11.15%) | 9 (5.5%) | 49 (13.1%) | 6 (14%) | 67 (11.15%) |
| Missing | 0 (0%) | 1 (0.7%) | 0 (0%) | 0 (0%) | 1 (0.2%) |
| **Patient income** |  |  |  |  |  |
| Less than 10 000\$ | 0 (0%) | 5 (3.6%) | 10 (2.7%) | 4 (9.3%) | 19 (3.3%) |
| 10 000 to 19 999\$ | 6 (23.1%) | 40 (29.0%) | 64 (17.1%) | 9 (20.9%) | 119 (20.5%) |
| 20 000 to 29 999\$ | 5 (19.2%) | 34 (24.6%) | 65 (17.4%) | 5 (11.6%) | 109 (18.8%) |
| 30 000 to 39 999\$ | 3 (11.5%) | 13 (9.4%) | 43 (11.5%) | 3 (7.0%) | 62 (10.7%) |
| 40 000 to 49 999\$ | 5 (19.2%) | 5 (3.6%) | 34 (9.1%) | 2 (4.7%) | 46 (7.9%) |
| 50 000 to 59 999\$ | 1 (3.8%) | 1 (0.7%) | 11 (2.9%) | 2 (4.7%) | 15 (2.6%) |
| 60 000 to 69 999\$ | 1 (3.8%) | 2 (1.4%) | 8 (2.1%) | 2 (4.7%) | 13 (2.2%) |
| 70 000 to 79 999\$ | 0 (0%) | 1 (0.7%) | 6 (1.6%) | 1 (2.3%) | 8 (1.4%) |
| 80 000 to 89 999\$ | 0 (0%) | 2 (1.4%) | 1 (0.3%) | 1 (2.3%) | 4 (0.7%) |
| 90 000 to 99 999\$ | 0 (0%) | 0 (0%) | 2 (0.5%) | 0 (0%) | 2 (0.3%) |
| More than 100 000\$ | 0 (0%) | 2 (1.4%) | 11 (2.9%) | 1 (2.3%) | 14 (2.4%) |
| Missing | 5 (19.2%) | 33 (23.9%) | 119 (31.8%) | 13 (30.2%) | 170 (29.3%) |
| **Patient housing type** |  |  |  |  |  |
| Home alone, intermediate or family-type residences, or public housing | 5 (11.5%) | 43 (29.0%) | 70 (18.2%) | 14 (32.6%) | 132 (21.5%) |
| Home, shared with a spouse or family | 19 (73.1%) | 80 (58.0%) | 267 (71.4%) | 25 (58.1%) | 391 (67.3%) |
| Retirement home | 2 (7.7%) | 15 (10.9%) | 37 (9.9%) | 4 (9.3%) | 58 (10.0%) |
| **Patient-Caregiver relationship** |  |  |  |  |  |
| Friend, sibling, or other | 4 (15.4%) | 11 (8.0%) | 45 (12.0%) | 6 (14.0%) | 66 (11.4%) |
| Parent-Child | 9 (34.6%) | 64 (46.4%) | 116 (31.0%) | 19 (44.2%) | 208 (35.8%) |
| Spouse | 13 (50.0%) | 63 (45.7%) | 212 (56.7%) | 18 (41.9%) | 306 (52.7%) |
| Missing | 0 (0%) | 0 (0%) | 1 (0.3%) | 0 (0%) | 1 (0.2%) |
| **Caregiver age** |  |  |  |  |  |
| Mean (SD) | 64.7 (11.2) | 62.8 (13.2) | 65.4 (11.8) | 62.0 (13.7) | 64.5 (12.3) |
| **Caregiver gender** |  |  |  |  |  |
| Man | 3 (11.5%) | 45 (32.6%) | 118 (31.6%) | 10 (23.3%) | 176 (30.3%) |
| Woman | 23 (88.5%) | 93 (67.4%) | 256 (68.4%) | 33 (76.7%) | 405 (69.7%) |
| **Caregiver first language** |  |  |  |  |  |
| French | 26 (100%) | 137 (99.3%) | 370 (98.7%) | 43 (100%) | 576 (99.0%) |
| English | 0 (0%) | 1 (0.7%) | 4 (1.1%) | 0 (0%) | 5 (0.9%) |
| **Caregiver highest level of education** |  |  |  |  |  |
| Primary school | 9 (34.6%) | 56 (40.6%) | 142 (38.0%) | 18 (41.9%) | 225 (38.7%) |
| High school (DES) | 6 (23.1%) | 45 (32.6%) | 98 (26.2%) | 11 (25.6%) | 160 (27.5%) |
| College (DEC) | 7 (26.9%) | 17 (12.3%) | 64 (17.1%) | 8 (18.6%) | 96 (16.5%) |
| Vocational studies (DEP) | 3 (11.5%) | 12 (8.7%) | 36 (9.6%) | 3 (7.0%) | 54 (9.3%) |
| University studies | 1 (3.8%) | 8 (5.8%) | 34 (9.1%) | 3 (7.0%) | 46 (7.9%) |
| Graduate school | 0 (0%) | 0 (0%) | 0 (0%) | 0 (0%) | 0 (0%) |
| **Caregiver income** |  |  |  |  |  |
| Less than 10 000\$ | 1 (3.8%) | 6 (4.3%) | 8 (2.1%) | 2 (4.7%) | 17 (2.9%) |
| 10 000 to 19 999\$ | 3 (11.5%) | 19 (13.8%) | 25 (6.7%) | 4 (9.3%) | 51 (8.8%) |
| 20 000 to 29 999\$ | 4 (15.4%) | 11 (8.0%) | 42 (11.2%) | 6 (14.0%) | 63 (10.8%) |
| 30 000 to 39 999\$ | 2 (7.7%) | 11 (8.0%) | 43 (11.5%) | 9 (20.9%) | 65 (11.2%) |
| 40 000 to 49 999\$ | 4 (15.4%) | 18 (13.0%) | 35 (9.4%) | 3 (7.0%) | 60 (10.3%) |
| 50 000 to 59 999\$ | 1 (3.8%) | 8 (5.8%) | 18 (4.8%) | 3 (7.0%) | 30 (5.2%) |
| 60 000 to 69 999\$ | 0 (0%) | 4 (2.9%) | 22 (5.9%) | 0 (0%) | 26 (4.5%) |
| 70 000 to 79 999\$ | 3 (11.5%) | 3 (2.2%) | 9 (2.4%) | 1 (2.3%) | 16 (2.8%) |
| 80 000 to 89 999\$ | 0 (0%) | 2 (1.4%) | 12 (3.2%) | 1 (2.3%) | 15 (2.6%) |
| 90 000 to 99 999\$ | 0 (0%) | 2 (1.4%) | 9 (2.4%) | 0 (0%) | 11 (1.9%) |
| More than 100 000\$ | 1 (3.8%) | 16 (11.6%) | 25 (6.7%) | 4 (9.3%) | 46 (7.9%) |
| Missing | 7 (26.9%) | 38 (27.5%) | 126 (33.7%) | 10 (23.3%) | 181 (31.2%) |
| **ZBI score** |  |  |  |  |  |
| Mean (SD) | 5.73 (5.42) | 10.5 (7.87) | 6.57 (7.02) | 7.65 (7.21) | 7.55 (7.36) |
| **Delay between ED index visit and caregiver recruitment (days)** |  |  |  |  |  |
| Mean (SD) | 24.8 (15.8) | 22.9 (19.1) | 27.9 (33.0) | 27.1 (17.4) | 26.5 (28.7) |

\* **Dates from the Institut national de santé publique du Québec (INSPQ):** Pre-pandemic (Before March 13th, 2020), Wave 1 (Between March 13th, 2020, to 11th of July 2020), Between the end of wave 1 and the beginning of wave 2 (12th of July 2020 to the 22nd of August 2020).

Our final coding scheme contained three main themes: *Care in the emergency department*, *Emergency Department Discharge* and *Capacity to live at home* (Figure 2). We counted when caregivers specifically mentioned each of three statements: an explicit call for help or information, a mention of time or financial costs associated with caregiving, and a mention of their CR’s autonomy (Figure 3). After making these amendments, 50 randomly selected comments were analyzed per coder, and again inter-rater reliability was very high (= .991).

**Figure 2.**
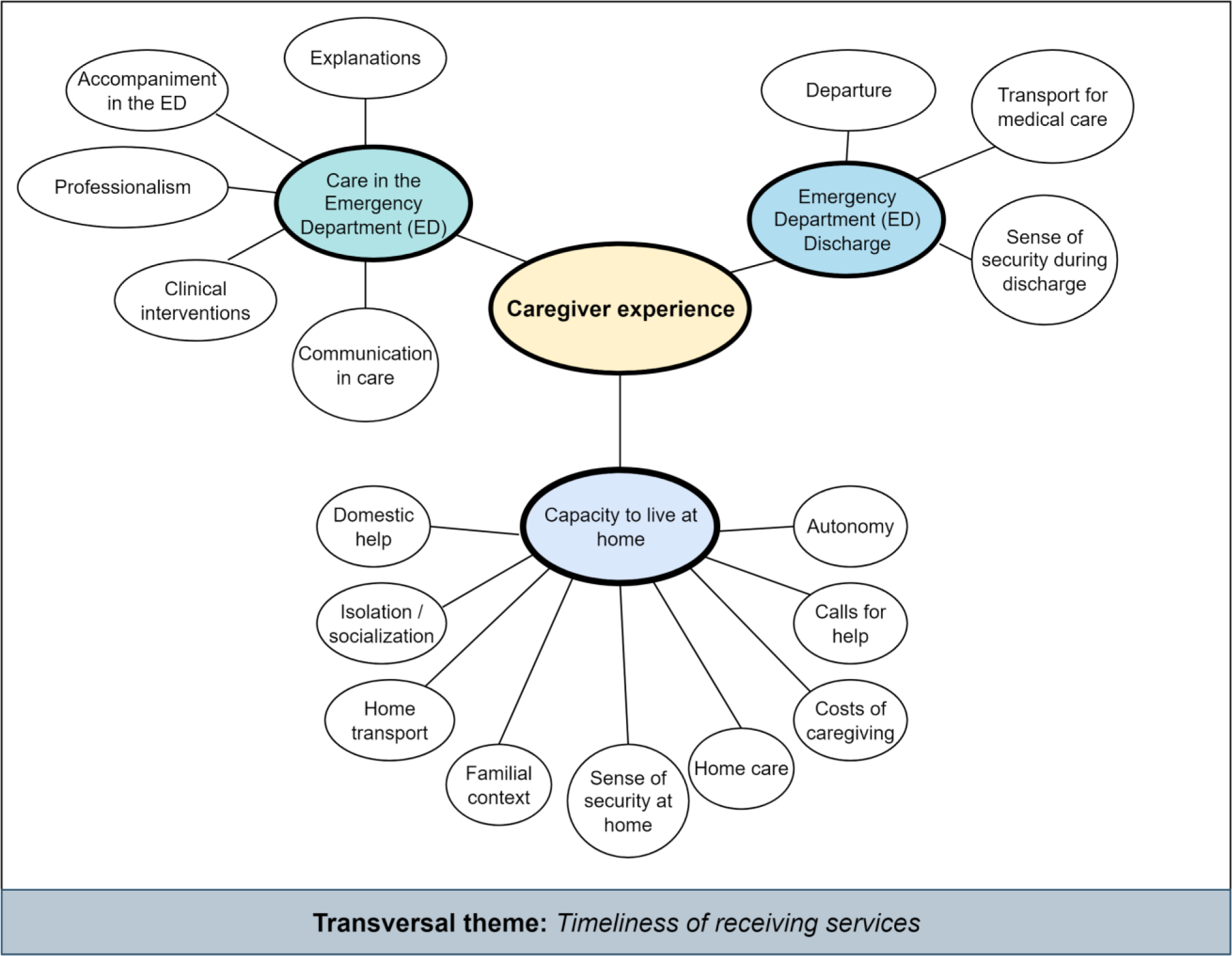
Mind map of the three main themes and eighteen sub-themes emerging from caregivers’ responses to the question: “In your opinion, what could be improved to reduce the burden of care for your [CR]?” following their CR’s transition of care from the emergency department to home. A transversal theme, *Timeliness of receiving services* is also identified. See Figure 3 for frequencies of each sub-theme mentioned and a graphical representation of the relative proportion of emotional valence associated with each theme.

**Figure 3.**
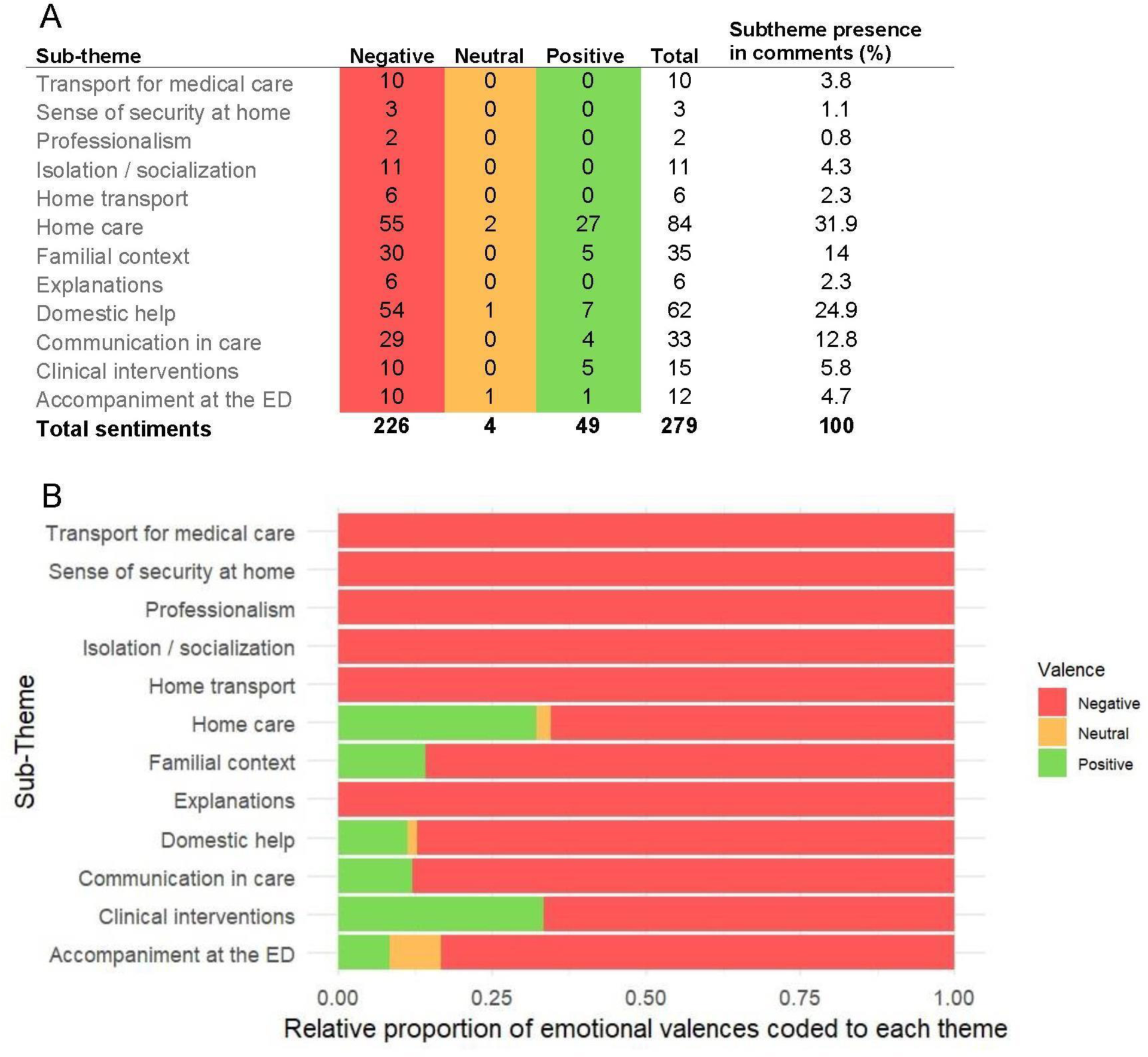
**A.** Frequency of sub-themes, stratified by emotional valence emerging from 253 caregiver comments. **B.** Relative proportions of negative, neutral, and positive sentiments coded to each theme embedded in 253 caregiver comments. For both A and B, red represents negative theme mentions, yellow neutral, and green positive.

**Figure 4.**
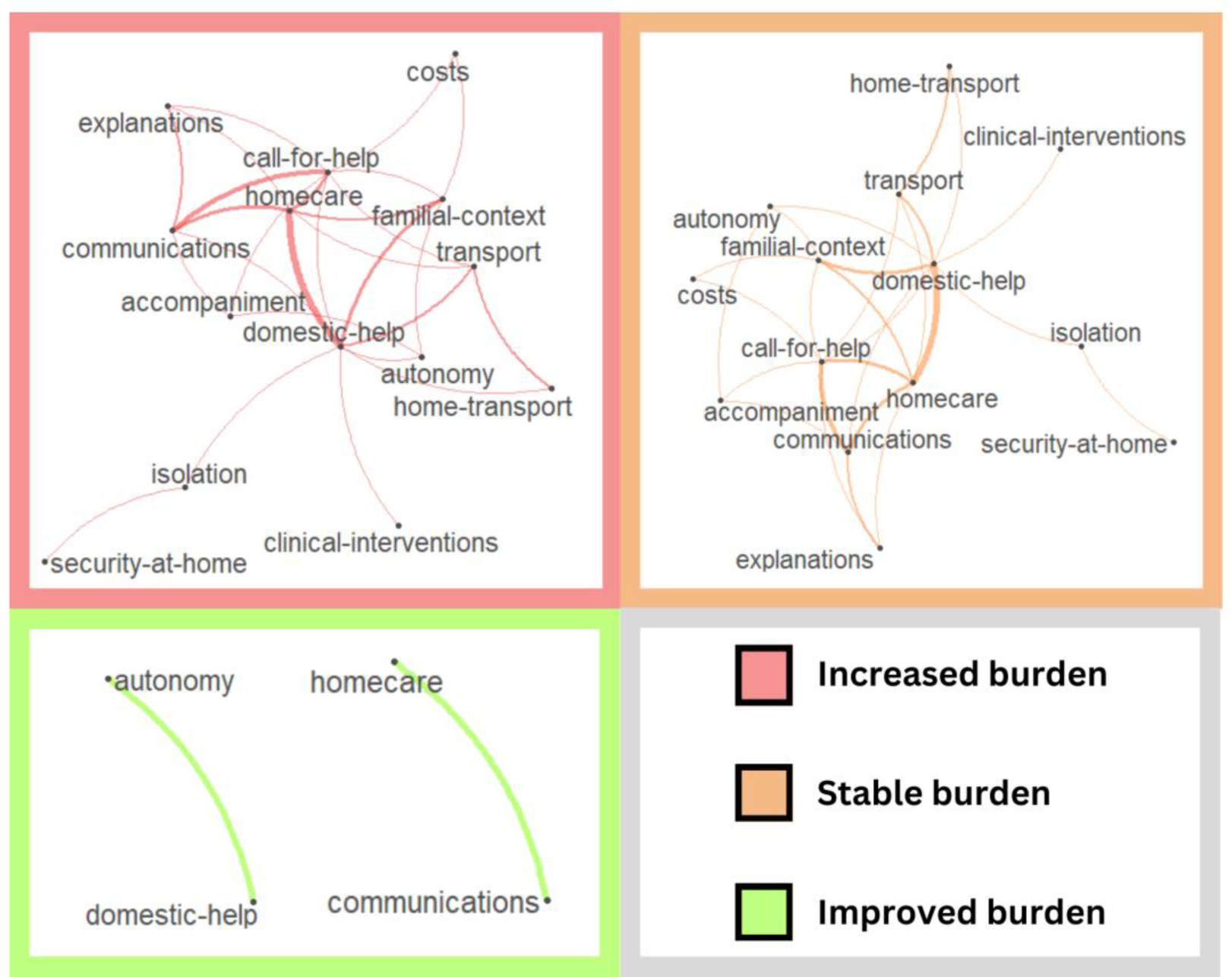
Co-occurrence networks of themes appearing in comment, by self-reported level of burden following departure from the emergency department. Lines between themes indicate co-occurrences within the same comment. Thinner lines represent fewer co-occurrences between two themes, and thicker lines denote more frequent co-occurrences.

#### Care in the emergency department

Both comments citing *Professionalism* in the emergency department were negative. One caregiver felt their physician was being negligent in their duties, changing the CR’s prescriptions without consulting the CR nor the caregiver. Another caregiver felt the personnel at the ED lacked humanity and understanding of the situation.

*Explanations* were viewed as negative by 6 caregivers. Two caregivers mentioned a lack of instruction for managing their CR’s conditions post discharge. One caregiver mentioned that they would have preferred to speak to the physician themselves because they felt as though their CR was withholding information and was being stubborn about adherence to treatment. Two caregivers mentioned having to relay information to the CR, who did not understand the information given to them by personnel. One caregiver mentioned that a physician changed their family member’s prescriptions without explanation.

Regarding *Communication in care*, 5 caregivers reported successful follow-up between departments or with specialists following their departure from the ED. Among negative commenters, 3 reported that they were still waiting to see a specialist, 8 mentioned gaps in access to information or wanting to know where and how to request different services. Four specifically suggested telephone follow-up calls to pass along updates and to see how CRs are managing at home. Four specifically mentioned difficulty reaching their family physician, and the remaining comments referred to waiting for the ED to transfer information or requests to other departments or outside services like convalescence homes, social services, and specialists.

Caregivers who reported *Accompaniment* of their CR to the ED tended to view this theme negatively. Five caregivers mentioned help with, or having someone else accompany their CRs to their appointments would be helpful. Four caregivers mentioned that accompanying their CR was personally taxing or affected their work schedule. One caregiver described relief that ten months of treatment requiring weekly visits was coming to an end. One caregiver reported positively, and one neutrally that they accompanied their CR to their medical appointments following an initial visit to the ED. One caregiver reported frustration that despite being there during the appointment, she was not allowed into the doctor’s office and that her mother would forget what the doctor had told her.

Of the 15 caregivers citing *Clinical intervention*s in their comments, 7 mentioned the quality of care, of which 2 were negative. One caregiver felt the management of their CR’s condition had room for improvement, while another caregiver felt that their CR was being treated with medications that only made them sicker, saying “*They just give her pills. She overdoses and then they give her other pills.*” The remaining comments cited care at the ED as good or excellent. Three other sub-themes emerged, and all were negatively referenced. One caregiver cited an error with medications, 3 felt the waiting time at the ED was too long, and one caregiver felt their CR was not given the correct diagnosis, saying “…*that he would have been properly diagnosed, I would have liked him to be seen by and re-evaluated in geriatrics.*”

#### Capacity to live at home

This main theme related to services empowering older adults to stay independent at home was dominated by comments about *Home care*. We coded comments as negative if a service was requested, but not yet delivered, in addition to negative experiences with homecare services that were delivered. Of these, 37 (22% positive) mentioned requesting or receiving care from a local community service center (CLSC), 10 (40% positive) mentioned receiving or requesting home visits from a physician or nurse. Nineteen mentioned living in or requesting adapted living environments (74% positive), and 8 mentioned they were waiting to be assigned a family physician; all of which were negative.

Eleven caregivers mentioned *Isolation* as an area for improvement. Three mentioned wishing their CRs lived closer, and the remainder mentioned wanting someone (a volunteer, other family member, or guardian) to spend some time with their CR, either to help with loneliness or boredom, or to accompany them to activities outside the home.

Similarly, *Home Transport* was used to code 6 mentions of non-medical transport. All comments requested access to transport for everyday needs like grocery shopping, and two caregivers suggested that access to this kind of transport would allow their CRs to independently run errands.

The *Familial context* emerged 35 times in the dataset, with most comments negative (86%). Two caregivers mentioned they did not mind taking care of their spouses, seeing it as part of their role. Three others mentioned other family members pitched in to help, dividing the workload. One of these caregivers mentioned ongoing discussions between their mother and the rest of the family, trying to convince their mother sell the family home and move to a more manageable residence. Similar discussions were reported negatively, with 4 caregivers mentioning the main problem was convincing their CR to accept their condition or to accept help. Five caregivers expressed frustration that they were not involved in shared decision-making about care and would have liked to be part of that process. The remainder expressed conflict or frustration with other family members, usually a desire for other family members to pull their weight or visit their CR from time to time. One caregiver mentioned wishing her son could trust that she is able to adequately care for his father.

Concerning *Domestic help*, seven positive comments mentioned that the caregiver felt housekeeping or cooking duties were adequately addressed. One neutral comment reported on housekeeping: “*I can’t wait for the person to come back!*”. Caregivers with negative comments (54) mentioned requesting help with household cleaning, making meals, and shoveling snow. Two caregivers mentioned a cooperative handled these tasks but canceled their services during the pandemic.

For three exploratory themes, we counted the occurrence but did not code for emotional valence, expecting each code to overlap somewhat with other codes (*Domestic help* and *Calls for help or information*, for example had considerable overlap; See co-occurrence networks section). *Calls for help or information* occurred 27 times, with caregivers requesting help for household tasks that they were not comfortable with (toileting, bathing). Most of these comments requested more services to help with respite, and to understand which services are available and how to request them through proper channels. One caregiver mentioned having information on how to “*get through it (or cope) when the situation becomes complicated*” would be helpful. Among comments citing the *Costs of caregiving*, 8 mentioned financial costs, with 2 caregivers mentioning that they would appreciate government financial aid. One caregiver mentioned an accumulation of stress that she felt was *costing* her own wellbeing. Caregivers who mentioned financial costs also often cited costs to time (4), mentioning taking time off work, or working less to fulfill their caregiving role. *Autonomy* was another common mention, occurring 35 times. In 28 cases, caregivers mentioned their CR was autonomous, and in these cases, caregivers reported overall positively. In 24 cases, there was nothing else to add in response. That is, autonomy was the only theme mentioned, and often in a one-word answer: “autonomous”. One caregiver reported that their CR was not autonomous, and that she dared not leave him alone. Another reported that their CR was living at home alone but suspected that they needed help with household tasks but would not ask for them.

#### Departure

We considered mentions of medical *Transport* to fall under a larger theme of *Departure* but all mentioned medical transport specifically: including transport to medical appointments and procedures (7 comments) or mentions that adapted transport should be improved to allow older CRs to autonomously attend their own appointments (3 comments). All these comments mentioned gaps in the availability of transport, so they were coded negatively.

#### Timeliness of services in and after the ED

The timeliness of receiving services in and after ED care was a transversal theme. We did not identify the timeliness of services as an individual sub-theme as it affected each major theme individually. For *Care in the emergency department*, 3 caregivers felt their wait time at the ED was too long, 3 caregivers mentioned long wait times to get follow-up care with a specialist after the ED visit, and 5 were still waiting for the ED to transfer information to a third party. For *Capacity to live at home*, issues with timeliness of services were most apparent, as we coded comments as negative if a service was requested but not yet delivered. The most frequent unmet requests for service were regarding awaiting services from community service centers (CLSCs; 29 mentions, including home care services specifically; 6 mentions), and waiting to be assigned a family physician (8 mentions). All comments mentioning *Discharge* were coded negatively: highlighting gaps in the availability of timely and convenient transport.

#### Caregivers who did not mention any theme

Of 331 caregivers who did not mention any themes in their comments, (85.7%) were neutral in tone, citing that they could not think of anything to report, or that they did not experience any burden of care to speak of. Forty-three caregivers mentioned that they had nothing to report because things were going well. One caregiver mentioned that things were going well and that they were content with the resources at their disposal. Another caregiver mentioned that everything was fine because she was retired and able to meet her CR’s needs, but acknowledged that if she was still working, things would be different. Only four caregivers left negative comments with no main themes emerging. One caregiver mentioned displeasure with their role as a caregiver, and another caregiver mentioned wishing that their CR would be able to function on their own. Another voiced that they could not think of anything that would help their CR, and a final comment mentioned their CR was in pain, but nothing could be done except to wait for the pain to pass.

#### Co-occurrence networks

Some caregivers gave comments containing more than one theme (*n* = 71). We stratified the dataset by self-reported change in burden (*increased* = 30, *stable* = 28, *improved* = 2; 11 caregivers did respond regarding a self-reported change in burden) to visualize which themes occur together and how they interact on this basis.

For caregivers reporting an unchanged (stable) burden following an ED care transition, central interconnected themes include communications, calls for help, home care, and domestic help. For caregivers reporting an increased burden, interconnected themes appear much more complex and include accompaniment, the familial context, costs, domestic help, explanations, calls for help, and communications. The bands linking domestic help to the familial context, costs, and calls for help are also thicker, indicating a greater importance of these themes in the comments of caregivers reporting an increase in subjective burden. For caregivers reporting a reduced burden, only two links emerged: the link between autonomy and domestic help, and homecare and communications. These links also emerge in the stable and increased burden groups, indicating that these two joined sets of themes may be important for all the caregivers we surveyed.

### Quantitative Results

#### Analysis of open-ended responses and the Zarit Burden Interview (ZBI)

The mean score on the ZBI among all caregivers included in the qualitative analysis (*N* = 581) was 7.55 (SD = 7.36), with a median of 5 (IQR = 2–11; Range = 0–38). The internal consistency was high (α = .879, 95% CI = [.86, .89])

#### Changes in caregiver burden following a visit to the emergency department

Cohen’s Kappa was calculated to quantify interrater reliability between the two coders (κ = 0.989, 647 comments). Results of the ANOVA analysis revealed statistically significant differences in the self-reported burden across the four categories (Binomial test χ^2^ = 530.5, p < .001): most caregivers reported that their level of burden did not change (64.3%), 23.8% of caregivers reported an increase in caregiver burden, 7.4% of caregivers reported a decrease in caregiver burden and 4.5% opted not to comment on the subject.

#### Changes in caregiver burden following a visit to the emergency department and scores on the ZBI

The effect of subjective change in burden on ZBI score was statistically significant (*F*(3, 191.62) = 11.83, *p* < .001). Caregivers who chose not to comment had the lowest ZBI scores (*n* = 26, *M* = 5.73, *SD* = 5.14), followed by caregivers with a level of burden left unchanged (*n* = 374, *M* = 6.56, *SD* = 7.02). Next were caregivers with a level of burden that improved following their CR’s visit to the emergency department (*n* = 43, *M* = 7.65, *SD* = 7.21). Caregivers who reported an increase in their subjective burden had the highest ZBI scores (*n* = 138, *M* = 10.53, *SD* = 7.87). ZBI scores among the increased burden group were statistically significantly different from the unchanged burden group (*t*(510) = 5.54, *p* < .001, *d* = 0.55), and the no comment group (*t*(162) = 3.12, *p* = .01, *d* = 0.66).

There was a statistically significant effect of change in burden on ZBI score even when controlling for age and comorbidities (*F*(3, 575) = 11.03, *p* < .001), and there was a statistically significant effect of Charlson Comorbidity Index score on ZBI score (*F*(1, 575) = 4.95, *p* = .026) but there was no effect of CR age on ZBI score (*F*(1, 575) = 0.007, *p* = .935).

## Discussion

We conducted a mixed methods design to understand, from the caregiver’s point of view, A) changes in burden of care following transitioning a CR’s care from the emergency room and B) what can be improved in the CR’s transition of care in this context. We also leveraged the French version of the Zarit Brief Burden interview (ZBI-12) to corroborate scores on caregiver burden with caregiving realities as reported by caregivers.

Changes in subjective burden appeared to correspond with ZBI scores. Greater ZBI scores are given to mean a greater level of caregiver burden. Most caregivers reported that their level of burden did not change (64.3%), and their average ZBI score was 6.57/48. Caregivers reporting an increase in burden (23.8%) had an average ZBI of 10.5. Mean scores of caregivers who reported an improvement in subjective burden (7.4%) were 7.65. Only the ZBI scores of the caregivers experiencing an increase in burden statistically differed from caregivers with stable burden. This effect remained when controlling for age and comorbidities of the CR. In the original ZBI-12 validated with caregivers of CRs with dementia, scores between 10 and 20 indicated moderate burden and scores greater than 20, high burden [37]. The cut-point signaling significant caregiver burden in other populations usually occurs in the teens and can be as low as 11[38], 12, [39], 13 [40], up to 17 [41,42]. This ZBI cutoff score appears to increase as the patient’s cognitive function decreases [42].

Caregivers who have higher levels of burden likely have greater room for improvements to burden, which may explain why caregivers in this improved burden group have higher ZBI scores than caregivers in the stable burden group. This effect has been documented in interventional studies aiming to reduce caregiver burden: caregivers with higher baseline burden experienced the greatest benefits in reducing burden [43]. Without baseline questionnaire data, we are limited to speculation as to how caregiver burden was affected by the care transition.

We also extracted themes from caregiver comments. Most caregivers did not mention any themes in their comments, most neutrally citing they had nothing to report. Caregivers providing comments reported concepts negatively, either indicating dissatisfaction or not yet receiving a service. *Home care* was the most prevalent sub-theme, with both positive and negative experiences discussed, followed by *Domestic help* and *Familial context*. Within home care related comments, outpatient care from community clinics and home visits from a physician or nurse were appreciated by caregivers. Depending on the chronicity of the illness, moving to, or suggesting an adapted living environment might be a way to reduce caregiver burden, according to caregivers. A referral to psychosocial or rehabilitation services like a psychologist, a social worker or an occupational therapist may be beneficial when discharging a patient from the ED.

While it may seem that these discussions may prove prohibitively time-consuming, one national American study found that three-quarters of primary care physicians felt responsible to identify caregiver needs when seeing patients, and half felt it important to address caregiver health and mental health in their assessment [44]. These physicians were four times as likely to take caregiver needs into consideration if they themselves acted in a caregiving role [44]. We are optimistic that while caregivers often cited gaps in home healthcare services, physicians appear to be open to integrate caregiver needs into care transition plans, especially as caregiving is becoming more common.

Communications and follow-up were also highlighted, mostly negatively. This theme has been negatively cited elsewhere in qualitative researchers of caregiver experiences in a transition from acute to community care [15,45–47]. Issues with the timeliness of services for home care were more pronounced in caregiver comments than by the patients in this same cohort [15]. This highlights the importance of involving caregivers in the construction of discharge plans as crucial to the success of care transitions. *Clinical interventions* were mentioned, with mixed sentiments, while *Accompaniment and Isolation* were predominantly viewed negatively. *Home Transport* and *Capacity to live at home* were discussed often with negative connotations. In a previous analysis of themes emerging from the patient in the care of these caregivers [15], patients were more likely to report on the quality of clinical interventions (their most common sub-theme). *Domestic help* was the most common sub-theme among caregivers, but *Communications and follow-up* were similarly referenced frequently as an area for improvement among both patients and their caregivers.

We split the dataset based on caregivers’ self-reported changes in burden (whether it increased, remained stable, or improved) to visually depict the co-occurrence and interactions of various sub-themes. Among caregivers who indicated a stable level of burden after an ED care transition, key interrelated sub-themes were *Communications*, *Calls for help*, *Home care*, and *Domestic help*. In cases of increased burden, the interconnections among sub-themes were more complex, and connections linking domestic help to family context, expenses, and appeals for assistance were more pronounced, indicating their heightened importance to this group. Caregivers reporting an improved burden only on *Autonomy* and *Domestic help*, and *Home care* and *Communications*, suggesting that these sub-themes are common to the experiences of many caregivers.

### Strengths and limitations

The strengths of our study arise from the applications of both quantitative and qualitative methodology, and the substantial random sample size at four different EDs. Our strong inter-rater reliability indicates a clear coding scheme, which we attribute to a rigorous, iterative development of an original coding scheme developed for patient comments.

Limitations of this study include the short nature of responses from caregivers. Open-response data sometimes lacks data substantial enough to achieve substantial credibility and resonance [48]. However, we were able to analyze several hundred comments—one way to boost the richness of otherwise sparse data. We acknowledge that an important proportion did not comment on their burden level (4.5 % replied “no comment” to Question A). The fact that caregivers took the time to complete the ZBI during this phone call, and provided generally short responses about improvements to care transitions might be a reflection of these caregivers truly not feeling or knowing what can be improved (48.7% of caregivers said they had nothing to report or nothing they feel could be improved in response to Question B) and not a result of insufficient sampling. We were also not able to distinguish between incident (sudden) and long-term caregivers [49] or describe caregivers’ own health status or comorbidities [50]. Both factors likely impact both caregiver burden and the quality of care transitions.

Another limitation is that comments for this study were collected over the telephone and transcribed immediately by a research professional. We did not audio record caregivers’ comments. This may have introduced an information bias such that the content has been filtered by the research professional conducting the interviews. Follow-up time poses another challenge, as patients were called to participate between 1–7 days following discharge. Follow-up time for caregivers (*M* = 26.52 days) was much more variable than the weeklong follow-up time of CRs. Questions posed soon after discharge may capitalize on the primacy of the caregiver’s experience but may not have left sufficient time to undergo all relevant aspects of the care transition, leading to an information bias.

## Conclusion

We used a mixed methods approach to understand the caregiver’s perspective regarding caregiver burden following a CR’s transition from the ED to home. To validate reported caregiver burden, we utilized the French version of the ZBI, comparing scores with caregivers’ real-world caregiving experiences. Only caregivers facing an increased self-reported burden showed significantly different ZBI scores compared to those with stable burden levels, which persisted even when accounting for CR age and comorbidities. This suggests that caregivers with greater initial burden may benefit most from targeted interventions designed to support caregiving. Findings can inform interventions aimed at reducing caregiver burden and enhancing the transition process, ultimately improving the well-being of both caregivers and those in their care.

## Data Availability

All anonymized data produced in the present study are available upon reasonable request to the authors.

## Declarations (unblinded)

### a. Ethics approval and consent to participate

The protocol for this study was approved by the CISSS-CA Ethics Review Committee (project #2018-462, 2018-007). Caregivers had to be able to understand French and provide informed consent. To demonstrate the capacity to provide informed consent, caregivers were required to summarize—in their own words—their understanding of the study to consent to, based on the Nova Scotia Criteria. Participants were also made aware that it would not be possible to identify individual participants in the published results.

### b. Consent for publication

Not applicable

### c. Availability of data and materials

The datasets created and analysed during the current study are available from the corresponding author on reasonable request.

### f. Authors’ contributions

1. Study concept and design
2. Acquisition of the data
3. Analysis and interpretation of the data
4. Drafting of the manuscript
5. Critical revision of the manuscript for important intellectual content
6. Statistical expertise
7. Obtained funding
8. Administrative, technical, or material support
9. Study supervision

1. NG, EJG, VC, MM, JR, ST, and PA conceptualized the study and analysis. NS and provided content expertise in study design and outcome selection.
2. ÉC, LA, RG, SC, ATF, JR, ST, RS, and PA collected and managed participant data.
3. NG, EJG, and PA conducted the analysis.
4. NG, and PA wrote the first draft of the manuscript.
5. SS, NS, LB, LBC, FL, HOW, and PA.

All authors critically reviewed the manuscript and provided comments to improve the manuscript. All authors read and approved the final manuscript.

6. RS, ST, LB, and FL provided statistical and methodological expertise.
7. FL, HOW, and PA obtained funding for this study.
8. ÉC, LA, RG, SC, ATF and JR, provided administrative and technical support.
9. PA is the principal investigator.

## g. Acknowledgements

We acknowledge the invaluable support of participating patients and their caregivers. We also thank Lise Lavoie, David Buckeridge, Josée Chouinard, Yves Couturier, Marie-Soleil Hardy, Audrey-Anne Brousseau, Éric Mercier, Clémence Dallaire, Richard Fleet, Annie Leblanc, Don Melady, Marie-Josée Sirois, Marcel Émond, Josée Rivard, Isabelle Pelletier and Jean-Louis Denis for their support and expertise in planning and contributing to the LEARNING WISDOM project.

## Network of Canadian Emergency Researchers

Patrick Archambault, MD, MSc^1,2,4,5^

## References

1. Rowe JW. The US eldercare workforce is falling further behind. Nat Aging 2021;1:327–9.

2. Stall N. We should care more about caregivers. CMAJ 2019;191:E245–6.

3. Williams A, Sethi B, Duggleby W et al. A Canadian qualitative study exploring the diversity of the experience of family caregivers of older adults with multiple chronic conditions using a social location perspective. Int J Equity Health 2016;15:40.

4. Brodaty H, Donkin M. Family caregivers of people with dementia. Dialogues Clin Neurosci 2009;11:217–28.

5. Schulz R, Sherwood PR. Physical and Mental Health Effects of Family Caregiving. Am J Nurs 2008;108:23–7.

6. Gabayan GZ, Asch SM, Hsia RY et al. Factors associated with short-term bounce-back admissions after emergency department discharge. Ann Emerg Med 2013;62:136–144.e1.

7. Hao S, Jin B, Shin AY et al. Risk Prediction of Emergency Department Revisit 30 Days Post Discharge: A Prospective Study. PLOS ONE 2014;9:e112944.

8. Mitchell SE, Laurens V, Weigel GM et al. Care Transitions From Patient and Caregiver Perspectives. Ann Fam Med 2018;16:225–31.

9. Rodakowski J, Rocco PB, Ortiz M et al. Caregiver Integration during Discharge Planning of Older Adults to Reduce Resource Utilization: A Systematic Review and Meta-Analysis of Randomized Controlled Trials. J Am Geriatr Soc 2017;65:1748–55.

10. Barnard D. What Is the Physician’s Responsibility to a Patient’s Family Caregiver? Commentary 1. AMA Journal of Ethics 2014;16:330–6.

11. Riffin C, Van Ness PH, Wolff JL et al. A Multifactorial Examination of Caregiver Burden in a National Sample of Family and Unpaid Caregivers. J Am Geriatr Soc 2019;67:277–83.

12. Riffina C, Wolff JL, Butterworth J et al. Challenges and Approaches to Involving Family Caregivers in Primary Care. Patient Educ Couns 2021;104:1644–51.

13. Laidsaar-Powell R, Butow P, Bu S et al. Family involvement in cancer treatment decision-making: A qualitative study of patient, family, and clinician attitudes and experiences. Patient Educ Couns 2016;99:1146–55.

14. Hébert R, Bravo G, Préville M. Reliability, validity and reference values of the Zarit Burden Interview for assessing informal caregivers of community-dwelling older persons with dementia. Canadian Journal on Aging 2000;19:494–507.

15. **[ANON]** Transitions of care for older adults discharged home from the emergency department: an inductive thematic content analysis of patient comments. BMC Geriatr 2024;24:8.

16. **[ANON]** Learning Integrated Health System to Mobilize Context-Adapted Knowledge With a Wiki Platform to Improve the Transitions of Frail Seniors From Hospitals and Emergency Departments to the Community (LEARNING WISDOM): Protocol for a Mixed-Methods Implementation Study. JMIR Res Protoc 2020;9:e17363.

17. O’Brien BC, Harris IB, Beckman TJ et al. Standards for reporting qualitative research: a synthesis of recommendations. Acad Med 2014;89:1245–51.

18. von Elm E, Altman DG, Egger M et al. The Strengthening the Reporting of Observational Studies in Epidemiology (STROBE) statement: guidelines for reporting observational studies. J Clin Epidemiol 2008;61:344–9.

19. Parry C, Mahoney EA, Chalmers, SA, et al. Assessing the quality of preparation for posthospital care from the patient’s perspective: the care transitions measure. Med Care 2005;46:317–322.

20. Kolk D, Kruiswijk AF, MacNeil-Vroomen JL et al. Older patients’ perspectives on factors contributing to frequent visits to the emergency department: a qualitative interview study. BMC Public Health 2021;21:1709.

21. College of Physicians and Surgeons of Nova Scotia. *Professional Standard and Guidelines Regarding Informed Patient Consent to Treatment*. Bedford, Nova Scotia, Canada, 2016.

22. Kühnel MB, Ramsenthaler C, Bausewein C et al. Validation of two short versions of the Zarit Burden Interview in the palliative care setting: a questionnaire to assess the burden of informal caregivers. Support Care Cancer 2020;28:5185–93.

23. Bianchi M, Flesch LD, Alves EV da C et al. Zarit Burden Interview Psychometric Indicators Applied in Older People Caregivers of Other Elderly. Rev Lat Am Enfermagem 2016;24:e2835.

24. Lu L, Wang L, Yang X et al. Zarit Caregiver Burden Interview: Development, reliability and validity of the Chinese version. Psychiatry and Clinical Neurosciences 2009;63:730–4.

25. Ozlu A, Yildiz M, Aker T. A Reliability and Validity Study on the Zarit Caregiver Burden Scale. Archives of Neuropsychiatry 2009;46.

26. Harris PA, Taylor R, Thielke R et al. Research electronic data capture (REDCap)--a metadata-driven methodology and workflow process for providing translational research informatics support. J Biomed Inform 2009;42:377–81.

27. Harris PA, Taylor R, Minor BL et al. The REDCap consortium: Building an international community of software platform partners. J Biomed Inform 2019;95:103208.

28. Chun Tie Y, Birks M, Francis K. Grounded theory research: A design framework for novice researchers. SAGE Open Med 2019;7:2050312118822927.

29. Elo S, Kyngäs H. The qualitative content analysis process. J Adv Nurs 2008;62:107–15.

30. Krippendorff K. *Content Analysis: An Introduction to Its Methodology*. SAGE Publications, Inc., 2019.

31. Krippendorff K. Computing Krippendorff’s Alpha-Reliability. Departmental Papers (ASC) 2011.

32. Archambault PM, Bilodeau A, Gagnon M-P et al. Health care professionals’ beliefs about using wiki-based reminders to promote best practices in trauma care. J Med Internet Res 2012;14:e49.

33. Turgeon AF, Dorrance K, Archambault P et al. Factors influencing decisions by critical care physicians to withdraw life-sustaining treatments in critically ill adult patients with severe traumatic brain injury. CMAJ 2019;191:E652–63.

34. Polit DF, Beck CT. *Nursing Research: Generating and Assessing Evidence for Nursing Practice*. Tenth edition. Philadelphia: Wolters Kluwer Health, 2017.

35. McHugh ML. Interrater reliability: the kappa statistic. Biochem Med (Zagreb) 2012;22:276– 82.

36. Benoit K, Watanabe K, Wang H et al. quanteda: An R package for the quantitative analysis of textual data. Journal of Open Source Software 2018;3:774.

37. Zarit SH, Reever KE, Bach-Peterson J. Relatives of the Impaired Elderly: Correlates of Feelings of Burden1. The Gerontologist 1980;20:649–55.

38. Hagell P, Alvariza A, Westergren A et al. Assessment of Burden Among Family Caregivers of People With Parkinson’s Disease Using the Zarit Burden Interview. J Pain Symptom Manage 2017;53:272–8.

39. Malik FA, Gysels M, Higginson IJ. Living with breathlessness: a survey of caregivers of breathless patients with lung cancer or heart failure. Palliat Med 2013;27:647–56.

40. Gratão ACM, Brigola AG, Ottaviani AC et al. Brief version of Zarit Burden Interview (ZBI) for burden assessment in older caregivers. Dement Neuropsychol 2019;13:122–9.

41. Bédard M, Molloy DW, Squire L et al. The Zarit Burden Interview: a new short version and screening version. Gerontologist 2001;41:652–7.

42. Stagg B, Larner A. Zarit Burden Interview: Pragmatic study in a dedicated cognitive function clinic. Progress in Neurology and Psychiatry 2015;19, DOI: 10.1002/pnp.390.

43. Towle RM, Low LL, Tan SB et al. Quality improvement study on early recognition and intervention of caregiver burden in a tertiary hospital. BMJ Open Qual 2020;9:e000873.

44. Park T, Pillemer K, Loeckenhoff C et al. What motivates physicians to address caregiver needs?: The role of experiential similarity. J Appl Gerontol 2023;42:1003–12.

45. Berntsen G, Høyem A, Lettrem I et al. A person-centered integrated care quality framework, based on a qualitative study of patients’ evaluation of care in light of chronic care ideals. BMC Health Serv Res 2018;18:479.

46. Boeykens D, Sirimsi MM, Timmermans L et al. How do people living with chronic conditions and their informal caregivers experience primary care? A phenomenological-hermeneutical study. Journal of Clinical Nursing 2023;32:422–37.

47. Pulcini CD, Belardo Z, Ketterer T et al. Improving emergency care for children with medical complexity: parent and physicians’ perspectives. Academic Pediatrics. 2021;21:513–520.

48. LaDonna KA, Taylor T, Lingard L. Why Open-Ended Survey Questions Are Unlikely to Support Rigorous Qualitative Insights. Academic Medicine 2018;93:347.

49. Liu C, Marino VR, Howard VJ et al. Positive aspects of caregiving in incident and long-term caregivers: Role of social engagement and distress. Aging & Mental Health 2023;27:87–93.

50. Kirvalidze M, Beridze G, Wimo A et al. Variability in perceived burden and health trajectories among older caregivers: a population-based study in Sweden. J Epidemiol Community Health 2023;77:125–32.

