## Supplemental Files: Appendices A to D for "Caregivers’ burden of care during emergency department care transitions among older adults: a mixed methods cohort study"

### Appendix A: Original open-ended questions (translated to English) and the Zarit Brief Burden Interview (English Translation of the Canadian French version)[1]

Question A:
*"In your opinion, has there been a change in the burden of care following your loved one's departure from the emergency department?"*

Question B:
*"In your opinion, what could be improved to reduce the burden of care for your loved one?"*

#### Zarit Brief Burden Interview (English Translation of the Canadian French version)

| | **Preamble: "How often do you…?"** *(____) refers to the name or relation (e.g., husband, sister) of the person being cared for.* |
|---|---|
| 1 | *Do you feel that because of the time you spend with (____) that you don't have enough time for yourself?* |
| 2 | *Do you feel stressed between caring for (____) and trying to meet other responsibilities for your family or work?* |
| 3 | *Worry about what the future holds for (____)?* |
| 4 | *Do you feel that (____) currently affects your relationships with other family members or friends in a negative way?* |
| 5 | *Do you feel strained when you are around (____)?* |
| 6 | *Do you feel your health has suffered because of your involvement with (____)?* |
| 7 | *Do you feel that you don't have as much privacy as you would like because of (____)?* |
| 8 | *Do you feel that your social life has suffered because you are caring for (____)?* |
| 9 | *Do you feel uncomfortable hosting friends because you are caring for (____)?* |
| 10 | *Do you feel you have lost control of your life since (____)'s illness?* |
| 11 | *Do you feel uncertain about what to do about (____)?* |
| 12 | *Overall, how burdened do you feel in caring for (____)?* |

Adapted from Hébert, R., Bravo, G., & Préville, M. (2000). Reliability, Validity and Reference Values of the Zarit Burden Interview for Assessing Informal Caregivers of Community-Dwelling Older Persons with Dementia. *Canadian Journal on Aging*, *19*(4), 494-507. https://doi.org/10.1017/S0714980800012484

[1] For French versions of the open-ended questions, ZBI questions, or French anonymized caregiver comments, please contact the corresponding author.

### Appendix B: Standards for Reporting Qualitative Research Checklist (SRQR)

**Page/line no(s).**

**Title and abstract**

| | |
|---|---|
| **Title** - Concise description of the nature and topic of the study Identifying the study as qualitative or indicating the approach (e.g., ethnography, grounded theory) or data collection methods (e.g., interview, focus group) is recommended | 1 |
| **Abstract** - Summary of key elements of the study using the abstract format of the intended publication; typically includes background, purpose, methods, results, and conclusions | 1 |

**Introduction**

| | |
|---|---|
| **Problem formulation** - Description and significance of the problem/phenomenon studied; review of relevant theory and empirical work; problem statement | 2 |
| **Purpose or research questio**n - Purpose of the study and specific objectives or questions | 2 |

**Methods**

| | |
|---|---|
| **Qualitative approach and research paradigm** - Qualitative approach (e.g., ethnography, grounded theory, case study, phenomenology, narrative research) and guiding theory if appropriate; identifying the research paradigm (e.g., postpositivist, constructivist/ interpretivist) is also recommended; rationale** | 3, 4 |
| **Researcher characteristics and reflexivity** - Researchers' characteristics that may influence the research, including personal attributes, qualifications/experience, relationship with participants, assumptions, and/or presuppositions; potential or actual interaction between researchers' characteristics and the research questions, approach, methods, results, and/or transferability | 5 |
| **Context** - Setting/site and salient contextual factors; rationale** | 3 |
| **Sampling strategy** - How and why research participants, documents, or events were selected; criteria for deciding when no further sampling was necessary (e.g., sampling saturation); rationale** | 5 |
| **Ethical issues pertaining to human subjects** - Documentation of approval by an appropriate ethics review board and participant consent, or explanation for lack thereof; other confidentiality and data security issues | 3 |

| | |
|---|---|
| **Data collection methods** - Types of data collected; details of data collection procedures including (as appropriate) start and stop dates of data collection and analysis, iterative process, triangulation of sources/methods, and modification of procedures in response to evolving study findings; rationale** | 3, 4 |
| **Data collection instruments and technologies** - Description of instruments (e.g., interview guides, questionnaires) and devices (e.g., audio recorders) used for data collection; if/how the instrument(s) changed over the course of the study | 4, 5, Appendix A |
| **Units of study** - Number and relevant characteristics of participants, documents, or events included in the study; level of participation (could be reported in results) | 6, 7, 8 |
| **Data processing** - Methods for processing data prior to and during analysis, including transcription, data entry, data management and security, verification of data integrity, data coding, and anonymization/de-identification of excerpts | Prior: 4<br>During: 5 |
| **Data analysis** - Process by which inferences, themes, etc., were identified and developed, including the researchers involved in data analysis; usually references a specific paradigm or approach; rationale** | 5 |
| **Techniques to enhance trustworthiness** - Techniques to enhance trustworthiness and credibility of data analysis (e.g., member checking, audit trail, triangulation); rationale** | 12: Reliability |

**Results/findings**

| | |
|---|---|
| **Synthesis and interpretation** - Main findings (e.g., interpretations, inferences, and themes); might include development of a theory or model, or integration with prior research or theory | 6-20 |
| **Links to empirical data** - Evidence (e.g., quotes, field notes, text excerpts, photographs) to substantiate analytic findings | 6-20 |

**Discussion**

| | |
|---|---|
| **Integration with prior work, implications, transferability, and contribution(s) to the field -** Short summary of main findings; explanation of how findings and conclusions connect to, support, elaborate on, or challenge conclusions of earlier scholarship; discussion of scope of application/generalizability; identification of unique contribution(s) to scholarship in a discipline or field | 21 |
| **Limitations** - Trustworthiness and limitations of findings | 22-23 |

**Other**

| | |
|---|---|
| **Conflicts of interest** - Potential sources of influence or perceived influence on study conduct and conclusions; how these were managed | Title document |
| **Funding** - Sources of funding and other support; role of funders in data collection, interpretation, and reporting | Title document |

*The authors created the SRQR by searching the literature to identify guidelines, reporting standards, and critical appraisal criteria for qualitative research; reviewing the reference lists of retrieved sources; and contacting experts to gain feedback. The SRQR aims to improve the transparency of all aspects of qualitative research by providing clear standards for reporting qualitative research.

**The rationale should briefly discuss the justification for choosing that theory, approach, method, or technique rather than other options available, the assumptions and limitations implicit in those choices, and how those choices influence study conclusions and transferability. As appropriate, the rationale for several items might be discussed together.

**Reference:**

O'Brien BC, Harris IB, Beckman TJ, Reed DA, Cook DA. **Standards for reporting qualitative research: a synthesis of recommendations.** *Academic Medicine*, Vol. 89, No. 9 / Sept 2014
DOI: 10.1097/ACM.0000000000000388

### Appendix C: STROBE Statement—Checklist of items that should be included in reports of *cohort studies*

| | Item No | Recommendation | Page No |
|---|---|---|---|
| **Title and abstract** | 1 | (*a*) Indicate the study's design with a commonly used term in the title or the abstract | Page 1. |
| | | (*b*) Provide in the abstract an informative and balanced summary of what was done and what was found | Page 1. |
| Introduction | | | |
| Background/rationale | 2 | Explain the scientific background and rationale for the investigation being reported | Pages 1 and 2. |
| Objectives | 3 | State specific objectives, including any prespecified hypotheses | Page 2, last paragraph |
| Methods | | | |
| Study design | 4 | Present key elements of study design early in the paper | First paragraph, methods section |
| Setting | 5 | Describe the setting, locations, and relevant dates, including periods of recruitment, exposure, follow-up, and data collection | Page 3. |
| Participants | 6 | (*a*) Give the eligibility criteria, and the sources and methods of selection of participants. Describe methods of follow-up | Page 3 and 4. |
| | | (*b*) For matched studies, give matching criteria and number of exposed and unexposed | |
| Variables | 7 | Clearly define all outcomes, exposures, predictors, potential confounders, and effect modifiers. Give diagnostic criteria, if applicable | Demographics in page 8. |

| | | | |
|---|---|---|---|
| Data sources/ measurement | 8* | For each variable of interest, give sources of data and details of methods of assessment (measurement). Describe comparability of assessment methods if there is more than one group | Pages 3 and 4. |
| Bias | 9 | Describe any efforts to address potential sources of bias | Reliability calculations (Page 5.)<br><br>Randomization and saturation (Page 5.) |
| Study size | 10 | Explain how the study size was arrived at | Page 5. Flowchart on page 7. |
| Quantitative variables | 11 | Explain how quantitative variables were handled in the analyses. If applicable, describe which groupings were chosen and why | Page 7. Page 5 describes the coding process. |
| Statistical methods | 12 | (*a*) Describe all statistical methods, including those used to control for confounding | Each analysis has its own subheading and description.<br><br>a. Reliability (Page 12), Frequency (Page 13), Narrative analysis (Pages 13-16), Co-occurrence networks (Page 18.), ZBI (Page 19.), Changes in burden, (Pages 19-20.). |
| | | (*b*) Describe any methods used to examine subgroups and interactions | b. Subroups split by emotional valence (Pages 13-14) and by changes in burden (Pages 19-20). |
| | | (*c*) Explain how missing data were addressed | c. Missing data were analyzed under caregivers who left no comment (Page 18.) |
| | | (*d*) If applicable, explain how loss to follow-up was addressed | d. N/A |

| | | | |
|---|---|---|---|
| | | (*e*) Describe any sensitivity analyses | e. N/A |
| Results | | | |
| Participants | 13* | (a) Report numbers of individuals at each stage of study—eg numbers potentially eligible, examined for eligibility, confirmed eligible, included in the study, completing follow-up, and analysed | Page 7. |
| | | (b) Give reasons for non-participation at each stage | Page 7. |
| | | (c) Consider use of a flow diagram | Page 7. |
| Descriptive data | 14* | (a) Give characteristics of study participants (eg demographic, clinical, social) and information on exposures and potential confounders | Table 1. Pages 8-11. |
| | | (b) Indicate number of participants with missing data for each variable of interest | Table 1. Pages 8-11. |
| | | (c) Summarise follow-up time (eg, average and total amount) | In Methods, Page 4. |
| Outcome data | 15* | Report numbers of outcome events or summary measures over time | Covid included as a covariate in Table 1. Page 9. |

| | | | |
|---|---|---|---|
| Main results | 16 | (*a*) Give unadjusted estimates and, if applicable, confounder-adjusted estimates and their precision (eg, 95% confidence interval). Make clear which confounders were adjusted for and why they were included | a. Not applicable. However, every quantitative analysis is presented with a measure of dispersion and its confidence interval.<br><br>b. Table 1. Pages 8-11.<br><br>c. N/A |

| | | | |
|---|---|---|---|
| | | (*b*) Report category boundaries when continuous variables were categorized | |
| | | (*c*) If relevant, consider translating estimates of relative risk into absolute risk for a meaningful time period | |
| Other analyses | 17 | Report other analyses done—eg analyses of subgroups and interactions, and sensitivity analyses | Pages 18-20 for non-strictly qualitative analyses. |
| Discussion | | | |
| Key results | 18 | Summarise key results with reference to study objectives | Page 21. |
| Limitations | 19 | Discuss limitations of the study, taking into account sources of potential bias or imprecision. Discuss both direction and magnitude of any potential bias | Page 21. |
| Interpretation | 20 | Give a cautious overall interpretation of results considering objectives, limitations, multiplicity of analyses, results from similar studies, and other relevant evidence | Page 22. |
| Generalisability | 21 | Discuss the generalisability (external validity) of the study results | Page 23. |
| Other information | | | |
| Funding | 22 | Give the source of funding and the role of the funders for the present study and, if applicable, for the original study on which the present article is based | Acknowledgements on page 23. |

*Give information separately for exposed and unexposed groups.

**Appendix D: Themes, selected caregiver comments, their translations, and definitions of each theme.**

| Theme | Example (translated) | Definition |
|---|---|---|
| Explanations | "It's difficult because I have a stubborn mother and I don't get to see the doctors myself. More complete explanations to help my mother would be appreciated because I feel like she's minimizing the problem to protect me. I'd like to make sure I help her properly." | Have received all relevant information delivered in an appropriate language to understand the interventions, results, and follow-up care in the emergency department. |
| Professionalism | "—they have good resources; you have to come across benevolent and understanding people." | Ability of ED staff to act professionally towards the patient. Reflects having respect, active listening skills, and compassion for the presenting patient. |
| Accompaniment to the ED | "More follow-up/ accompaniment outside the emergency. This could prevent him from returning so often for the same thing." | Have a familiar and trusted person accompany the patient during the stay at the ED and during the transition of care. |
| Communications in care | "Not much [can be done for] his condition, [it is] not something that can be easily dealt with. Well followed [by the care team], [there] is always someone who calls him if there is something." | Transfer of the patient's medical information (family history, test results, level of care, etc.) to all relevant stakeholders within an acceptable time frame. |
| Clinical interventions | "Currently very happy to have had treatment at [the hospital] (with rheumatology and internist). Good place to get treatment (specialists and multi-[disciplinary] team). This lightened their burden not long after the consultation." | Specific medical care and techniques performed in the hospital. |
| Sense of security at home | "[Would like] more security" | Ability to feel sheltered from danger, confident, safe, and at peace in one's living environment. |

| | | |
|---|---|---|
| Home care | "Would appreciate having a family doctor. We've been on the waiting list for two years. It's worrying." | Intervention or service performed outside of a hospital center (home, community clinic, pharmacy, etc.) which requires specific training or accreditation. For instance, home visits by a nurse, physician, social worker were considered part of home care. So too were appointments with a nutritionist, optometrist, occupational therapist, and consultations with a community pharmacist. |
| Isolation / Socialization | "There is nothing that can help [the patient] in the context of COVID-19. But in another context, she would need someone available to spend time with her. They certainly have some services like housekeeping and outdoor maintenance, and the physiotherapist, but what is really missing is someone to keep her company." | Ability to have support from a social network or another person. |
| Domestic help | "Get some outside help. For example, it was snowing this week, but [the patient] cannot shovel, so she had to drive 20 km to clear his driveway. Help with housekeeping would be nice too." | Someone who can offer help with daily chores and household tasks. |
| Home transport | "Have more volunteers available to take her [the patient] downtown (which she [the patient] likes to do but her caregiver doesn't like to do)." | Access to transportation for activities of daily living. |
| Familial context | "Involving more family members would be very helpful. [With] the availability to meet [the] needs [of the patient], since he [the caregiver] is the only one involved." | References the social and relational environment within a family, including individual or shared experiences. |
| Costs of caregiving | "I [the caregiver] am working 1 day less. [I wish I had] more time, and government funding for caregivers." | References to the fiscal or time costs associated with caregiving. |

| | | |
|---|---|---|
| Calls for help | “[Would like] better collaboration such that she would be able to know which services she is entitled to in order to get help (requests for help care like CLSC, help with showering).” | Refers to an explicit call for help or information from the commenting caregiver. |
| Autonomy | He [the patient] is autonomous, occasionally he has a small problem, but it is transient; it does not last long.” | References the autonomy or degree of dependence of the patient in the care of the caregiver. |
| Sense of security during discharge | No comments mentioned this theme | Feeling safe from danger while returning to their home living environment. |
| Transport for medical care | “[Would like ameliorations in] transportation so that she [the patient] can go to her appointments and do her errands by herself. Since my mother no longer has a car, I dedicate one day a week to travel so that she can buy her things and go to her [medical] appointments.” | Ability to have transportation or a vehicle for attending appointments related to their medical condition. |
| Departure | No comments mentioned this theme | Circumstance surrounding the conclusion of hospital care and the return to the patient’s usual living environment. |

(Note: These quotes are the written notes taken by the research professionals doing the phone interviews and the third person point of view is sometimes used when describing what the patient had said. The first-person point of view is used when the research professional is quoting the patient’s own words.)
